# Development and Validation of a Low-Cost 3DPrinted Cervical Simulator for Fluoroscopy-Guided Nerve Block Training

**DOI:** 10.64898/2025.12.10.25341989

**Authors:** Tomas Gondra, Romina Gimbatti

## Abstract

Cervical nerve blocks require high precision due to the proximity of critical structures, such as the spinal cord and the vertebral and carotid arteries. Training opportunities are limited by the risks associated with fluoroscopy-guided procedures, ethical constraints of cadaveric models, and the high cost of commercial simulators. Three-dimensional (3D) printing offers an accessible and customizable alternative for procedural training. A cervical spine simulator was constructed from a multislice CT scan. The cervical spine (C1-C7) was segmented using 3D Slicer and processed in Meshmixer to generate an anatomical 3D model, which was printed in PLA. Soft tissues were reproduced using a ballistic gel formulation composed of glycerin, gelatin, and propylene glycol. Six physicians participated in a five-week training program performing facet, indirect, and direct cervical root blocks under fluoroscopy. Knowledge was assessed pre- and post-training, performance was assessed weekly using OSATS, and validity was evaluated through a Likert-scale survey. This 3D-printed cervical simulator proved to be a valid, low-cost, and reproducible tool for training fluoroscopy-guided cervical nerve blocks.

## INTRODUCTION

Cervical radicular pain affects approximately 0.1% of the population per year. When cervical radiculopathy becomes symptomatic, brachial pain is present in 99% of cases and can reach an intensity that significantly impacts quality of life and work performance. In the majority of cases, appropriate pain management is achieved with conventional medical treatment. When medical therapy fails, minimally invasive procedures, such as nerve blocks, can provide symptom relief. Currently, these procedures are performed with increasing frequency at the lumbar level but far less commonly at the cervical level. This is mainly due to the complexity of cervical anatomy, which includes vital structures such as the spinal cord, vertebral artery, carotid artery, and jugular vein. Their proximity requires a high degree of technical precision [1-4]. In an environment where patient safety is paramount, simulation and deliberate practice have become essential pillars of training for both residents and specialists. The limitations of traditional learning in the operating room have encouraged training programs to adopt simulation-based workshops and laboratory sessions. The goal of simulation is to provide a safe learning environment that allows trial and error without posing risks to patients or participants. The use of cadaveric or animal material presents logistical, ethical, maintenance, and availability challenges, and commercial simulators-although useful-are often inaccessible due to their high cost [5-7].

Advances in 3D-printing technology have enabled the creation of low-cost, reproducible anatomical simulators suitable for safe training in a controlled environment. The present study evaluates the usefulness of a low-cost 3D-printed cervical simulator designed for the practice of facet and nerve root blocks.

## TECHNICAL REPORT

### Simulator fabrication

To create the simulator, a multislice computed tomography scan of the cervical spine in DICOM format was used as the starting point. The bony segment of interest-specifically the cervical vertebrae from C1 to C7-was segmented using the open-source software 3D Slicer 5.8.1, generating a three-dimensional model that was subsequently exported in STL format.

The resulting file was edited and refined in Meshmixer 3.5.474, where anatomical details were adjusted, and the final geometry was prepared for printing. The bony structure was printed in polylactic acid (PLA) using a home-grade FDM 3D printer (Weedo F152s). This material was selected based on its low cost and appropriate behavior under fluoroscopy. Print parameters were configured using Ultimaker Cura 5.10.

**Figure 1.**
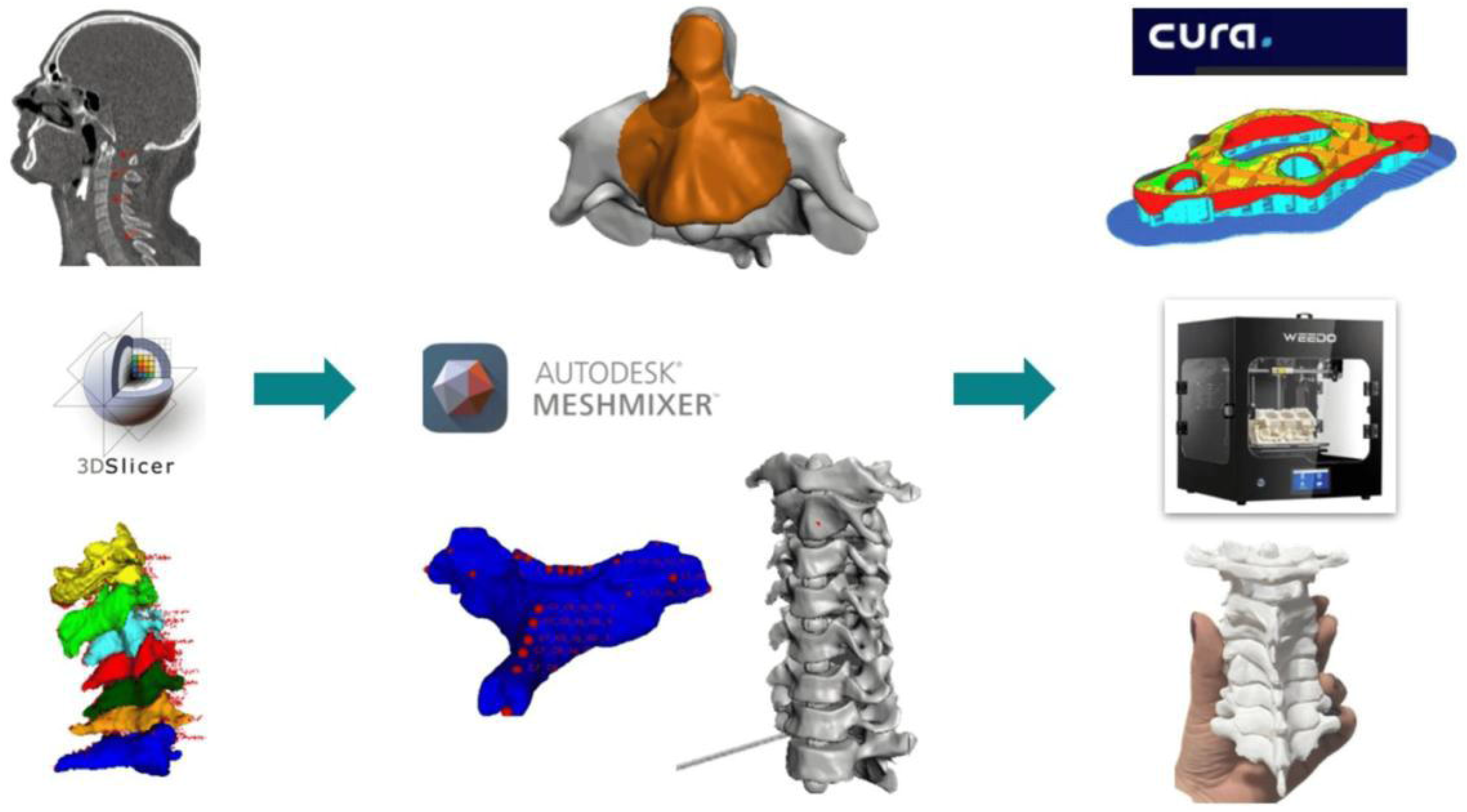

The surrounding soft tissues were produced using a ballistic gel composed of water, glycerin, Bloom 280 gelatin, propylene glycol, and acrylic pigment. The material was poured into molds created in Blender 4.1 and subsequently printed using flexible TPU 95 filaments. Different proportions of these components were tested to reproduce the texture, resistance, and mechanical response of real tissues during needle manipulation. Once all components were completed, the simulator was assembled onto a support structure also designed in Blender and printed in PLA, providing stability during training maneuvers. The total cost of the simulator was 40 USD.

**Figure 2.**
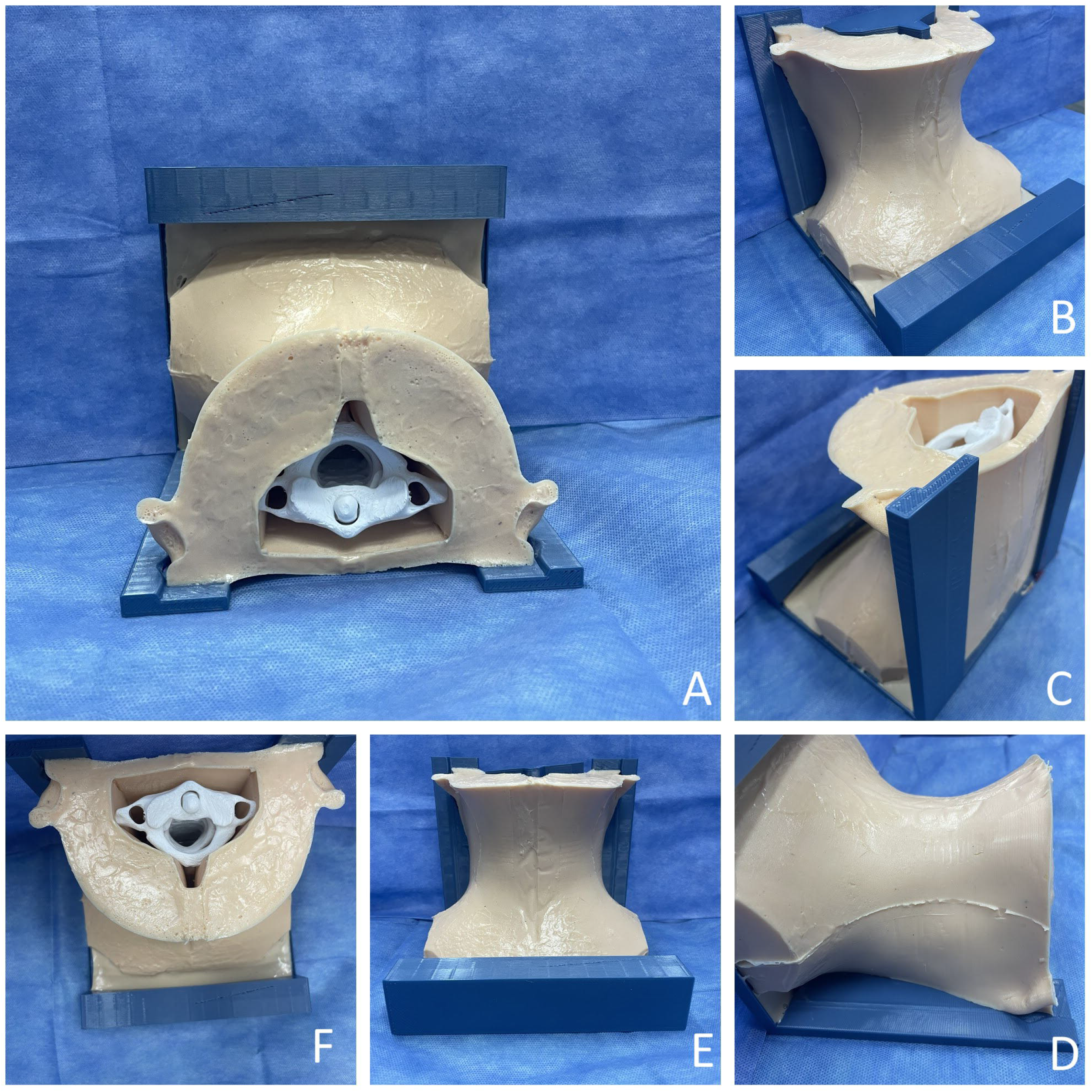

**Figure 3.**
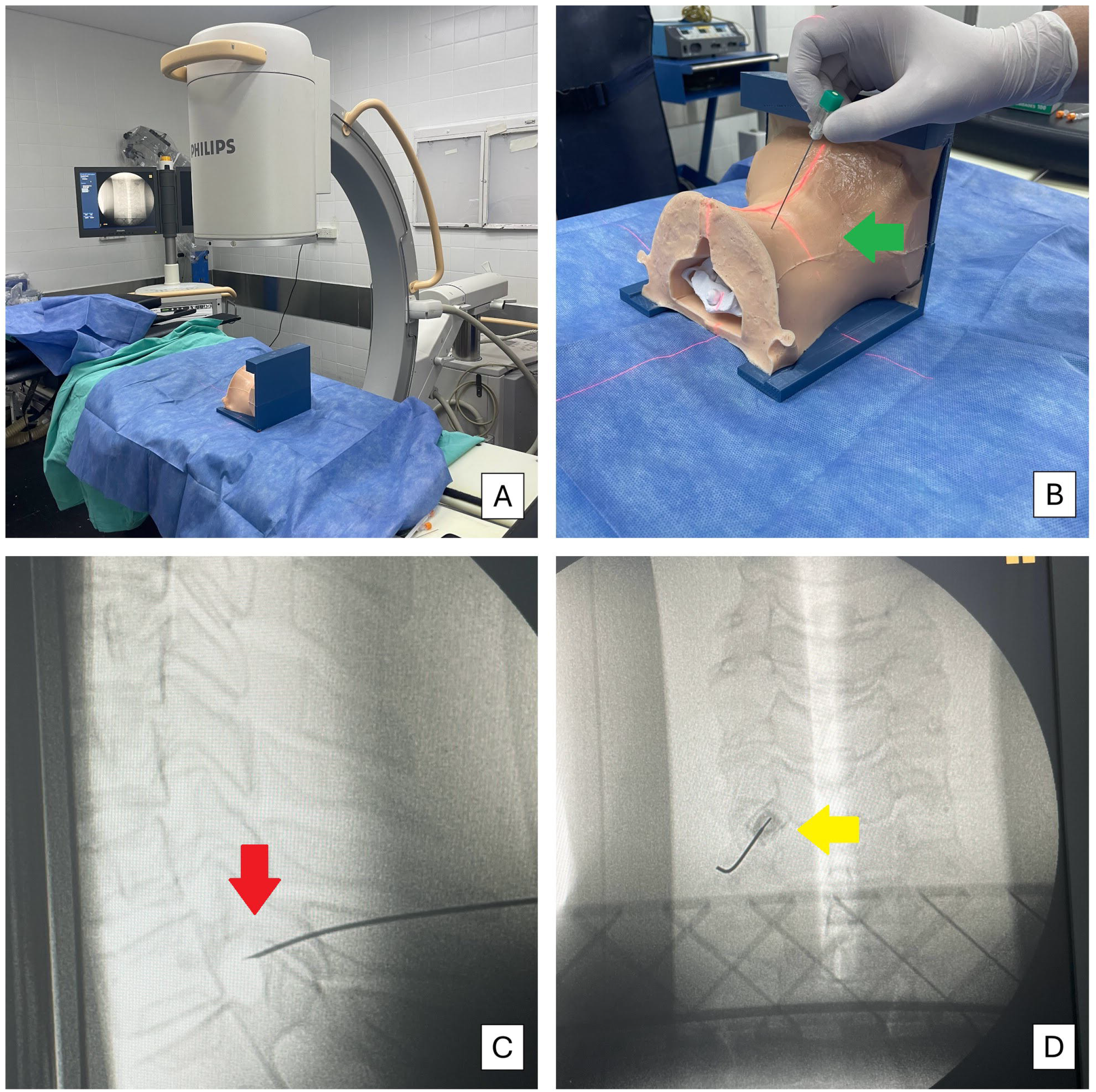

### Participants

Six physicians were included in the study (four residents and two junior neurosurgeons). The resident group was designated as group A, and the specialist group as group B. None of the participants had prior formal experience with fluoroscopy-guided cervical blocks. All volunteers agreed to participate in the educational program.

### Training protocol

The training program was conducted over five consecutive weeks, with one session per week for each participant. In every session, the physicians performed two attempts for each of the evaluated techniques: facet block, indirect nerve root block, and direct nerve root block. The procedures were performed under fluoroscopic guidance using a BV Endura Phillips unit, with a 21-G spinal needle. The training environment was designed to closely replicate typical operating-room conditions, including simulator positioning in the prone orientation, fluoroscopic identification of bony structures, and needle advancement under continuous imaging guidance. The program included an assessment of theoretical knowledge, practical skills, and simulator validity.

### Theoretical learning assessment

Before and after the training program, participants completed a test focused on the recognition of anatomical structures on fluoroscopic images, with a maximum score of 5 points. The images used included anteroposterior, lateral, and oblique projections of the cervical spine, requiring identification of anatomical landmarks relevant to the procedure (Table 1).

**TABLE 1:**
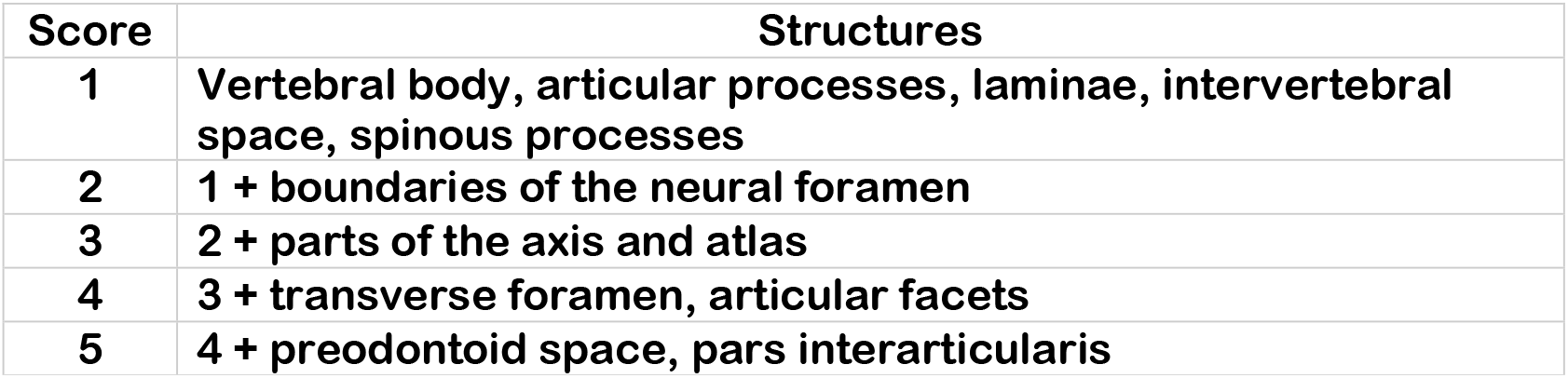
Anatomical structures and scoring criteria. This scoring system is not derived from previously published classification systems (e.g., SINS, SLIC, AOSpine) and does not represent a validated or standardized framework in the medical literature. Instead, the table was designed by the authors to organize anatomical structures in a progressive manner for educational and procedural assessment purposes.

| Score | Structures |
| --- | --- |
| 1 | Vertebral body, articular processes, laminae, intervertebral space, spinous processes |
| 2 | 1 + boundaries of the neural foramen |
| 3 | 2 + parts of the axis and atlas |
| 4 | 3 + transverse foramen, articular facets |
| 5 | 4 + preodontoid space, pars interarticularis |

After the training program, an improvement in anatomical recognition was observed. In Group A, the mean score increased from 2.5 points (range 1-4) at the beginning of the program to 4.5 points (range 4-5) at the end. No changes were observed in Group B.

### Practical performance assessment

Technical performance was evaluated using an instrument based on the Objective Structured Assessment of Technical Skills (OSATS), modified for spinal procedures. The performance rating scale used in this study was adapted from the global rating scale originally developed for the Objective Structured Assessment of Technical Skill (OSATS) by Martin et al. (1997). This framework employs a five-point scale anchored at scores 1, 3, and 5 with behavioural descriptors, while intermediate values (2 and 4) represent performance levels between adjacent anchors. Consistent with this methodology, we preserved the anchored descriptors and maintained intermediate scores without specific definitions, as these serve to capture gradations of performance without imposing artificial categorical distinctions. [8] The assessment was conducted by a neurosurgeon with experience in spinal interventional techniques (Table 2). All participants demonstrated improvement on the OSATS, with an average score of 3.7 in the first week and 4.8 in the final week of training. (Table 3)

**TABLE 2:** Performance rating scale adapted from the global rating scale described by Martin et al for the OSATS model. [8] **Scores 1, 3, and 5 provide anchor descriptors of performance quality. Scores 2 and 4 represent intermediate performance levels between adjacent anchors and were intentionally left without specific descriptors** This scoring structure follows the validated OSATS global rating approach, which anchors scores 1, 3, and 5 with explicit descriptors while allowing 2 and 4 to function as intermediate, non-anchored performance levels.

| Skill | Score 1 | Score 2 | Score 3 | Score 4 | Score 5 |
| --- | --- | --- | --- | --- | --- |
| **Movement effectiveness** | Insecure, inefficient, no progression | Intermediate | Slow, reasoned, organized | Intermediate | Confident, efficient, fluid |
| **Needle handling** | Misses target, slow correction | Intermediate | Some target errors but quick correction | Intermediate | Precise direction, correct plane, minimal readjustments |
| **Minimizes simulator damage** | Rough, tears material, little control | Intermediate | Minor trauma with occasional damage | Intermediate | Appropriate tension, minimal damage |
| **Procedure flow** | Insecure, constantly shifting focus | Intermediate | Slow but reasonably planned and organized | Intermediate | Confident, maintains focus and advances |

**TABLE 3:**
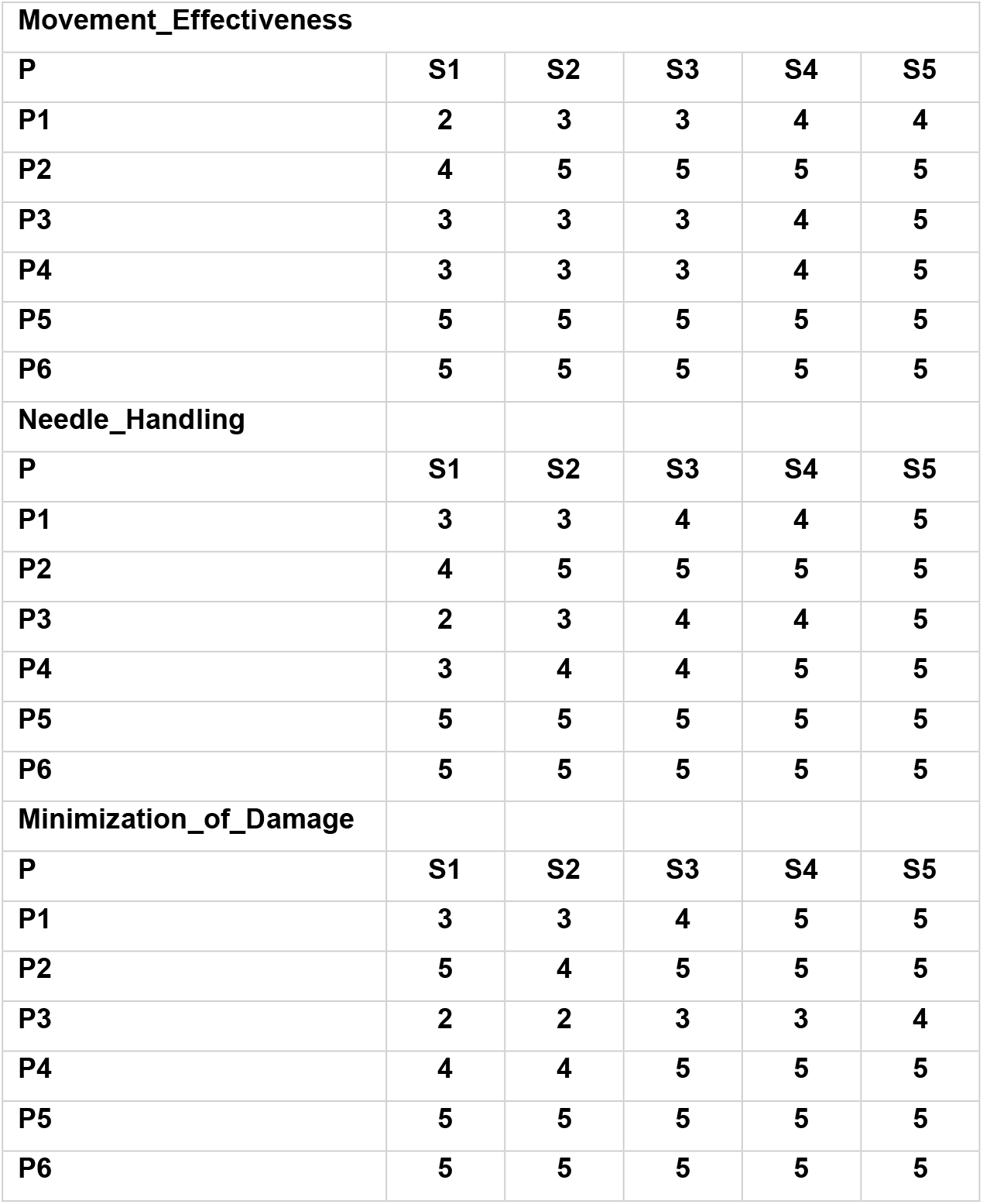
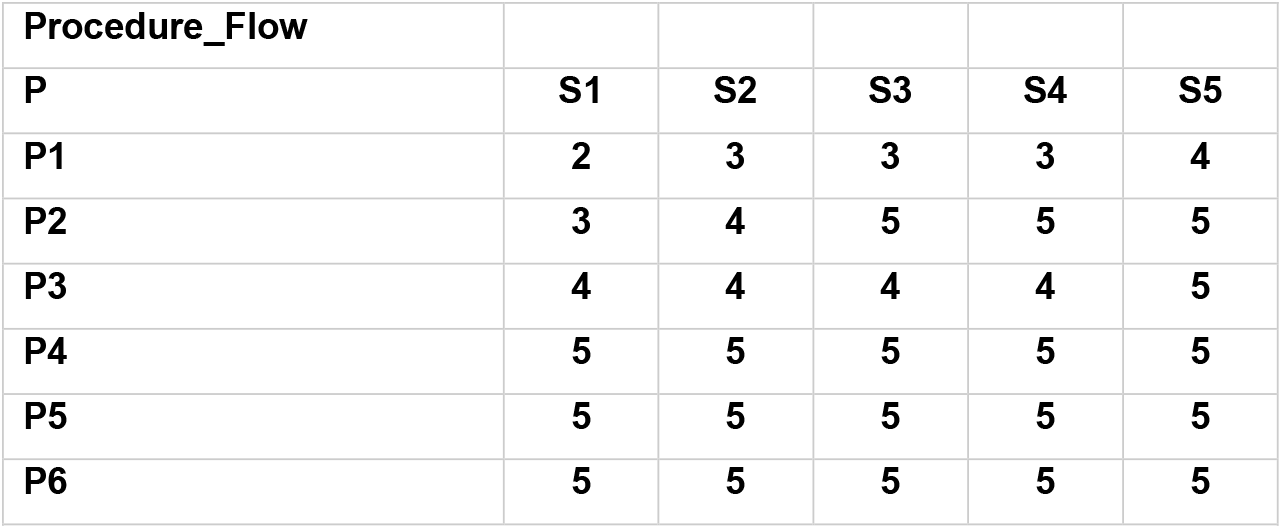
Evolution of Technical Performance: OSATS Scores Across Four Domains Over Five Training Sessions. P: participant / S: session Scores correspond to all participants (P1–P6) evaluated across five consecutive weekly sessions (S1–S5) using the adapted OSATS global rating framework.

### Simulator Validation

To evaluate the simulator’s validity, all the participants and a spine specialist completed a 5-point Likert scale assessing the following domains: anatomical fidelity, fluoroscopic visualization, material elasticity, usefulness for learning and practice, aesthetic quality, likelihood of recommending the simulator to a colleague, and overall rating. In the Likert-scale evaluation (n = 7), most participants rated the simulator’s aesthetics as “excellent” in 4 (57%), the anatomical structural fidelity as “excellent” in 5 (71%), and the fluoroscopic visualization as “excellent” in 6 (86%). The usefulness for practice was rated as “very useful” by 5 (71%) and as “useful” by 2 (29%). Regarding material elasticity, 4 (57%) rated it as “intermediate,” 2 (29%) as “elastic,” and 1 (14%) as “very elastic”. A total of 5 (71%) agreed that the model facilitated learning of the technique, and 6 (86%) reported that they would recommend the simulator. The overall rating was ≥8 points in 7 (100%) of cases: 3 (43%) scored 8 points, 1 (14%) scored 9 points, and 3 (43%) scored 10 points. (Table 4)

**TABLE 4:**
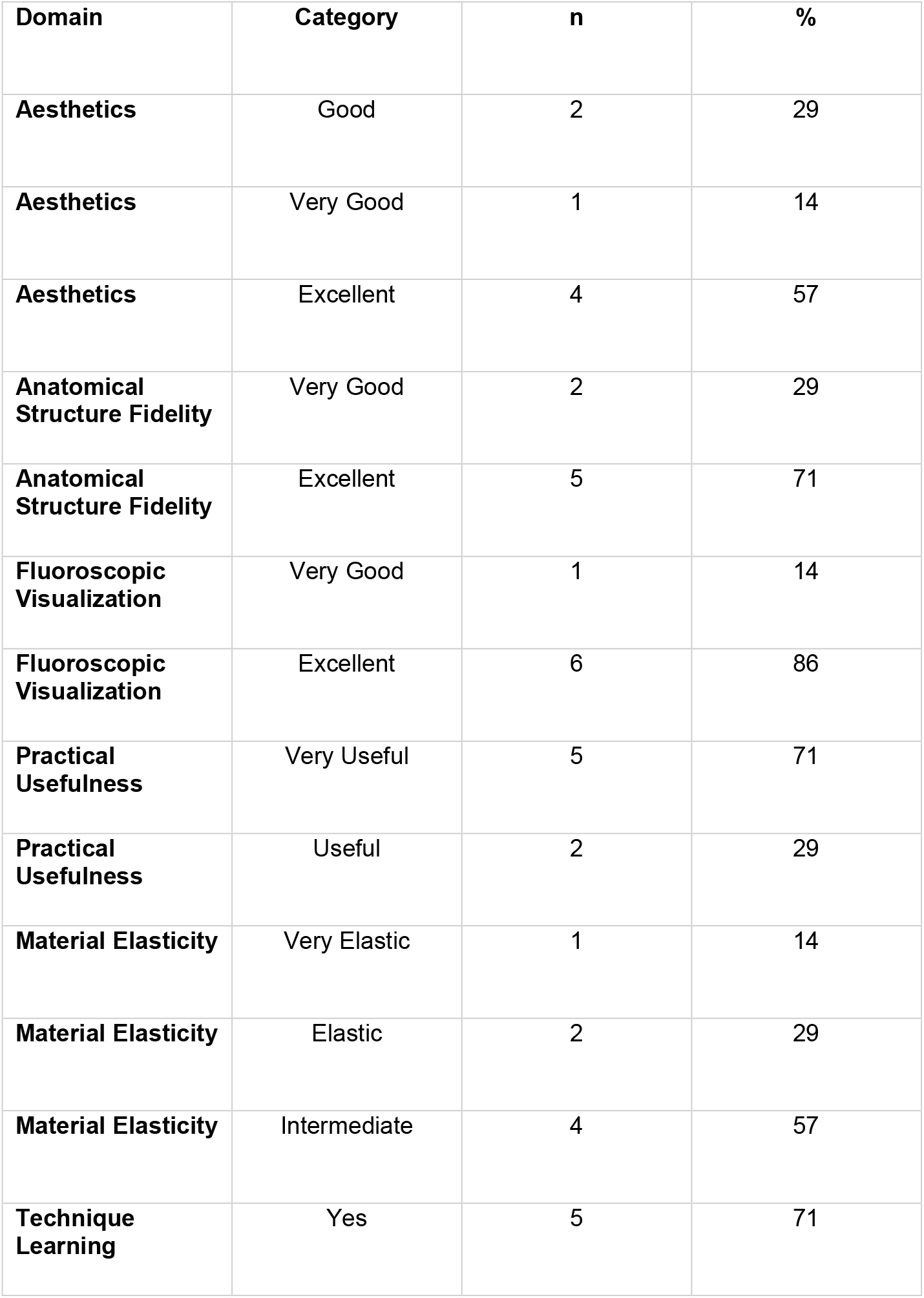
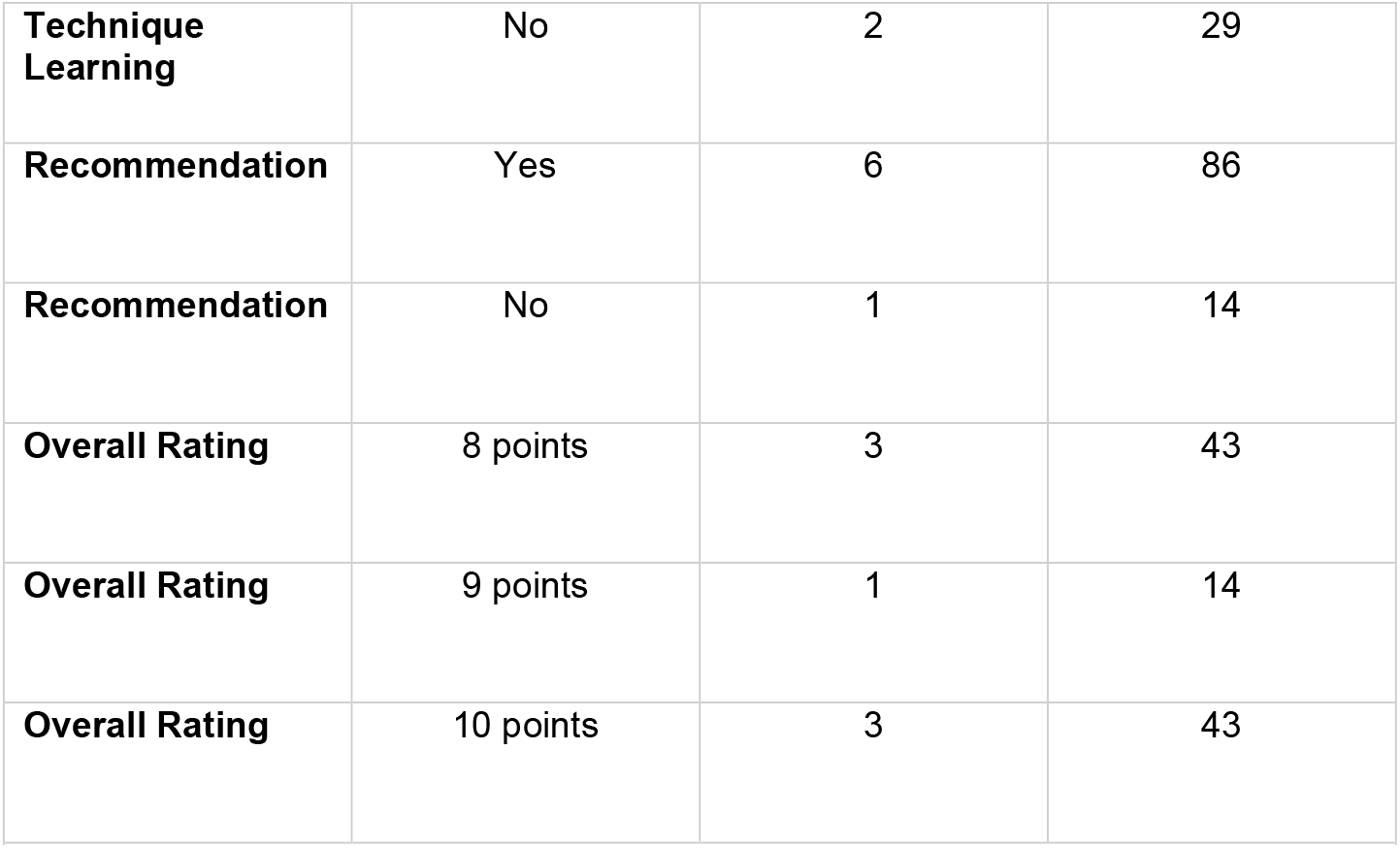
Results of the simulator validation survey completed by all participants (n = 7). Values are expressed as n (%).

| **Domain** | **Category** | **n** | **%** |
| --- | --- | --- | --- |
| **Aesthetics** | Good | 2 | 29 |
| **Aesthetics** | Very Good | 1 | 14 |
| **Aesthetics** | Excellent | 4 | 57 |
| **Anatomical Structure Fidelity** | Very Good | 2 | 29 |
| **Anatomical Structure Fidelity** | Excellent | 5 | 71 |
| **Fluoroscopic Visualization** | Very Good | 1 | 14 |
| **Fluoroscopic Visualization** | Excellent | 6 | 86 |
| **Practical Usefulness** | Very Useful | 5 | 71 |
| **Practical Usefulness** | Useful | 2 | 29 |
| **Material Elasticity** | Very Elastic | 1 | 14 |
| **Material Elasticity** | Elastic | 2 | 29 |
| **Material Elasticity** | Intermediate | 4 | 57 |
| **Technique Learning** | Yes | 5 | 71 |
| **Technique Learning** | No | 2 | 29 |
| **Recommendation** | Yes | 6 | 86 |
| **Recommendation** | No | 1 | 14 |
| **Overall Rating** | 8 points | 3 | 43 |
| **Overall Rating** | 9 points | 1 | 14 |
| **Overall Rating** | 10 points | 3 | 43 |

### Limitations

This study has several limitations that should be considered when interpreting the results. First, the number of participants was small, as is common in early-phase feasibility studies, which limits the generalizability of the findings. Given this small sample size, the analysis was intentionally restricted to descriptive statistics. Inferential statistical testing (e.g., ANOVA, t-tests, chi-square) was not performed because the sample size did not meet the assumptions required for these methods and could lead to misleading or uninterpretable results. Second, the assessment relied on a single expert evaluator, which may introduce observer bias despite the use of a structured and validated rating framework. Finally, the study evaluated short-term performance trends across consecutive training sessions but did not assess long-term skill retention or transferability to real clinical settings. Future work with larger cohorts, multiple evaluators, and longitudinal follow-up will be essential to further validate the educational impact of this simulator. Despite these limitations, the findings support the feasibility and educational value of the proposed cervical spine simulator.

## DISCUSSION

The acquisition of perceptual, psychomotor, and spatial-orientation skills is fundamental for performing fluoroscopy-guided interventional procedures. These skills rely not only on accurate interpretation of imaging but also on the coordinated manipulation of needles, guidewires, and other instruments [9,10]. Because trainees generally exhibit longer fluoroscopy times and higher radiation exposure during early learning, practicing in a safe environment prior to patient contact is essential [11]. Simulation-based training has therefore emerged as a key strategy to improve procedural competency while reducing risks to patients. Existing literature supports the role of simulation in accelerating skill acquisition. Patel et al. reported significant improvements in proficiency across multiple interventional disciplines following simulationbased programs [12]. Phantom models in particular have demonstrated educational value, especially among beginners, by enabling repetition and feedback without compromising safety. Although cadaveric and animal models offer high tactile realism, their increasing cost, limited accessibility, and ethical concerns have restricted their widespread use [13,14]. Meanwhile, virtual reality and commercial simulators provide sophisticated training options but remain financially prohibitive for many centers. In this context, digital manufacturing and 3D printing have gained prominence as practical alternatives for producing anatomically accurate, customizable, and cost-effective training tools. The use of patient-derived DICOM data to generate three-dimensional models has been applied successfully across multiple surgical specialties for preoperative planning and skill development [14-16]. Such technology offers the possibility of democratizing access to simulation resources, particularly in environments where high-cost systems are unattainable. The validity of a simulator depends on its ability to realistically replicate procedural conditions and measurably improve targeted skills [17, 18]. The cervical spine model developed in this study was designed to meet these criteria by providing fluoroscopic realism, anatomical fidelity, and appropriate resistance during needle manipulation. Through a structured training curriculum, participants demonstrated measurable improvement in both anatomical recognition and technical performance, suggesting effective early-phase skill acquisition. Additionally, subjective evaluations showed high acceptance regarding realism, usefulness, and likelihood of recommending the simulator to peers. Taken together, these findings indicate that a low-cost, 3D-printed cervical phantom can serve as an accessible and pedagogically valuable alternative to traditional high-fidelity models. Its affordability and reproducibility make it particularly suitable for resource-limited institutions, supporting broader dissemination of structured training in cervical spine interventions.

## CONCLUSIONS

This study demonstrates that a low-cost, 3D-printed cervical spine simulator can serve as an accessible and effective tool for developing fluoroscopy-guided percutaneous skills. The model provided adequate anatomical fidelity, realistic fluoroscopic behavior, and appropriate mechanical resistance, allowing trainees to practice needle manipulation in a safe and controlled environment. Participants showed progressive improvement in both anatomical recognition and technical performance, and subjective evaluations reflected high levels of acceptance regarding the simulator’s realism and educational usefulness. Given its affordability, reproducibility, and ease of implementation, this simulator represents a valuable training resource-particularly for programs with limited access to high-cost or cadaveric models-and may contribute to safer and more standardized pathways for early procedural learning. Future studies with larger cohorts and long-term follow-up are warranted to further assess its impact on clinical performance.

## Data Availability

All data produced in the present study are available upon reasonable request to the authors

## Additional Information

### Author Contributions

All authors have reviewed the final version to be published and agreed to be accountable for all aspects of the work.

### Concept and design

Tomas Gondra Sr., Romina Gimbatti

### Acquisition, analysis, or interpretation of data

Tomas Gondra Sr., Romina Gimbatti Drafting of the manuscript: Tomas Gondra Sr.

### Critical review of the manuscript for important intellectual content

Romina Gimbatti

## Disclosures

### Human subjects

All authors have confirmed that this study did not involve human participants or tissue.

### Animal subjects

All authors have confirmed that this study did not involve animal subjects or tissue.

## REFERENCES

1. Bono CM, Ghiselli G, Gilbert TJ, et al.: Evidence-based clinical guideline for the diagnosis and treatment of cervical radiculopathy from degenerative disorders. Spine J. 2011, 11(1):64–72. 10.1016/j.spinee.2010.10.023.

2. Radhakrishnan K, Litchy WJ, O’Fallon WM, Kurland LT: Epidemiology of cervical radiculopathy. Apopulation-based study from Rochester, Minnesota, 1976 through 1990. Brain. 1994, 117(2):325-335. 10.1093/brain/117.2.325

3. Woods BI, Hilibrand AS: Cervical radiculopathy: epidemiology, etiology, diagnosis, and treatment . J Spinal Disord Tech. 2015, 28(5):E251–E259. 10.1097/BSD.0000000000000284

4. Taso M, Sommernes JH, Kolstad F, et al.: A randomised controlled trial comparing the effectiveness of surgical and nonsurgical treatment for cervical radiculopathy. BMC Musculoskelet Disord. 2020, 21:171. 10.1186/s12891-020-3188-6

5. Byvaltsev VA, Belykh EG, Konovalov NA: New simulation technologies in neurosurgery . Zh Vopr NeirokhirIm N N Burdenko. 2016, 80(2):102–107. 10.17116/neiro2016802102-107.

6. Davids J, Manivannan S, Darzi A, Giannarou S, Ashrafian H, Marcus HJ: Simulation for skills training in neurosurgery: a systematic review, meta-analysis, and analysis of progressive scholarly acceptance. Neurosurg Rev. 2021, 44:1853–1867. 10.1007/s10143-020-01378-0

7. Alaraj A, Luciano CJ, Bailey DP, et al.: Virtual reality cerebral aneurysm clipping simulation with real-time haptic feedback. Neurosurgery. 2015, 11(Suppl 2):52–58. 10.1227/NEU.0000000000000583

8. Martin JA, Regehr G, Reznick R, MacRae H, Murnaghan J, Hutchison C, Brown M: Objective structured assessment of technical skill (OSATS) for surgical residents. Br J Surg. 1997, 84:273–278. 10.1046/j.1365-2168.1997.02502.x

9. Suri: Ashish; Patra, Devi Prasad; Meena, Rajesh Kumar. Simulation in neurosurgery: Past, present, and future. Neurology India. 2016, 64(3):387–395. 10.4103/0028-3886.181556

10. Gould D: Using simulation for interventional radiology training . Br J Radiol. 2010, 83:546-553. 10.1259/bjr/33259594

11. Dawson DL, Meyer J, Lee ES, Pevec WC: Training with simulation improves residents’ endovascularprocedure skills. J Vasc Surg. 2007, 45:149–154. 10.1016/j.jvs.2006.09.003

12. Patel R, Dennick R: Simulation based teaching in interventional radiology training: is it effective? . ClinRadiol. 2017, 72(3):266.e7-266.e14. 10.1016/j.crad.2016.10.014

13. Kim YH: Ultrasound Phantoms to Protect Patients from Novices. Korean J Pain. 2016, 29:73-77. 10.3344/kjp.2016.29.2.73.

14. Miller ZA, Amin A, Tu J, et al.: Simulation-based Training for Interventional Radiology and Opportunitiesfor Improving the Educational Paradigm. Tech Vasc Interv Radiol. 2019, 22 (1):35–40. 10.1053/j.tvir.2018.10.008

15. Javan R, Ellenbogen AL, Greek N, Haji-Momenian S: A prototype assembled 3D-printed phantom of theglenohumeral joint for fluoroscopic-guided shoulder arthrography. Skeletal Radiol. 2019, 48:791–802. 10.1007/s00256-018-2979-4.

16. Jahnke P, Schwarz FB, Ziegert M, et al.: A radiopaque 3D printed, anthropomorphic phantom for simulationof CT-guided procedures. Eur Radiol. 2018, 28:4818–4823. 10.1007/s00330-018-5481-4

17. Stunt J, Wulms P, Kerkhoffs G, Dankelman J, van Dijk C, Tuijthof G: How valid are commercially available medical simulators?. Adv Med Educ Pract. 2014, 5:385–95. 10.2147/AMEP.S63435

18. Carlotta Fontana, Emilio Cataldo, Giovanni Torelli, et al.: Validation of a virtual reality-based surgical training for pedicle screw placemen

